# The Visual Hemofilter: a novel visualization technology that improves task performance among intensive care professionals: A prospective simulation study

**DOI:** 10.64898/2026.04.16.26351012

**Authors:** Justyna Bider-Lunkiewicz, Greta Gasciauskaite, Blanca Rück Perez, Julia Braun, Jan Willms, Hunor Szekessy, Christoph B. Nöthiger, Matthias Hoffmann, Petar Milovanovic, Emanuela Keller, David W. Tscholl

## Abstract

**Purpose:** This study evaluates the Visual Hemofilter, a novel decision-support and information transfer tool designed to assist with regional citrate anticoagulation (RCA) in hemofiltration. By representing hemofilter parameters and patient blood constituents as animated icons, the tool aims to improve clinicians’ interpretation of blood gas results and RCA reference tables. We hypothesized that the Visual Hemofilter would enhance clinical decision-making by enabling faster and more accurate therapy adjustments, increasing clinicians’ confidence in their decisions, and reducing cognitive workload compared to conventional methods.

**Methods:** We conducted a prospective, randomized, computer-based simulation study across four intensive care units at the University Hospital Zurich. Twenty-six critical care professionals participated, each managing regional citrate anticoagulation (RCA) scenarios using either the Visual Hemofilter or conventional methods involving blood gas analysis and reference tables. Following each scenario, participants made therapy adjustments and rated their decision confidence and cognitive workload.

**Results:** Use of the Visual Hemofilter significantly improved decision accuracy (odds ratio [OR] 3.96; 95% CI 2.03–7.73; p < 0.0001) and reduced decision time by an average of 33 seconds (mean difference –33.3 seconds; 95% CI –39.4 to –27.2; p < 0.0001). Participants also reported greater confidence in their decisions (OR 5.41; 95% CI 2.49–11.77; p < 0.0001) and experienced lower cognitive workload (mean difference –15.05 points on the NASA-TLX scale (National Aeronautics and Space Administration-Task Load Index); 95% CI –18.99 to –11.13; p < 0.0001).

**Conclusions:** The Visual Hemofilter enhances clinical decision-making in RCA by increasing accuracy and speed, boosting decision confidence, and reducing cognitive workload. This technology has the potential to reduce errors and better support critical care professionals in managing complex treatment scenarios.

## Introduction

Situation awareness–oriented, human-centered visualization technologies in medicine leverage dynamic visual elements—such as shape, color, and motion—to support perceptual processing and foster a clearer, more intuitive understanding of the patient’s physiological state [1, 2]. These visualization approaches have been associated with improved recognition of clinical problems, enhanced diagnostic accuracy, and faster decision-making in time-critical scenarios [2–5].

The Visual Hemofilter (VHF) is a novel visualization and decision-support tool developed to assist with regional citrate anticoagulation (RCA) during hemofiltration—a recommended modality of continuous renal replacement therapy (CRRT) for hemodynamically unstable patients with severe acute kidney injury (AKI) [6–8]. While RCA has been shown to reduce complication rates compared to heparin [7, 9–12], it is also associated with a higher incidence of metabolic disturbances, particularly acid–base imbalances [6, 13, 14]. Many of these complications could be mitigated through timely therapy adjustments [6, 7, 15], highlighting the potential benefit of a dedicated visualization and decision-support tool. Time management is another significant challenge associated with RCA. Ongoing RCA management requires frequent blood gas analyses, parameter adjustments, and continuous system monitoring—making it a time-intensive process that imposes a substantial cognitive load on clinical staff. Studies have reported that intensive care nurses spend approximately eight minutes per hour managing CRRT alone [16]. Easily accessible data, presented in a single location and in a user-centered, intuitive format, could support more time-efficient decision-making [1]. To date, no visualization technologies exist to support critical care staff in monitoring regional citrate anticoagulation, detecting potential complications, and guiding clinical decision-making.

The Visual Hemofilter displays hemofilter parameters and patient blood components as animated icons flowing through a virtual filter. Its design incorporates user-centered design principles described by Mica Endsley [17] and is grounded in the principle that a model should exhibit logical commonality with the reality it represents [18], as well as insights from research on other medical visualization technologies [1]. When a specific parameter falls below a predefined threshold, the corresponding icons become dashed, fade, or blink. Conversely, when a parameter exceeds the threshold, the icons appear in greater quantity to signal abnormality. For example, blood flow is represented by animated erythrocytes; if the flow is too high, the animation depicts erythrocytes moving rapidly through the filter, prompting the user to reduce the rate to the target level. Dialysate flow is visualized as clear droplets falling at variable rates—too slow, normal, or too fast—depending on the actual flow. To reflect physiological interactions, the Visual Hemofilter also illustrates citrate binding to calcium ions, displays the rates of citrate and calcium infusion, and shows both systemic and post-filter calcium concentrations. Acid–base status is represented by varying quantities of hydrogen and bicarbonate ions. In the presence of acid–base imbalances, the VHF provides visual cues and suggests appropriate RCA adjustments to support the restoration of homeostasis.

Fig 1 presents representative screenshots of the Visual Hemofilter animation. A detailed description of the Visual Hemofilter is available in the instructional video provided in S1 Appendix. For this study, the VHF operated using an algorithm based on the citrate-calcium (CiCa) system (Fresenius, Bad Homburg vor der Höhe, Germany), and the findings specifically apply to the citrate-based continuous veno-venous hemodialysis modality (CiCa-CVVHD).

**Fig 1.**
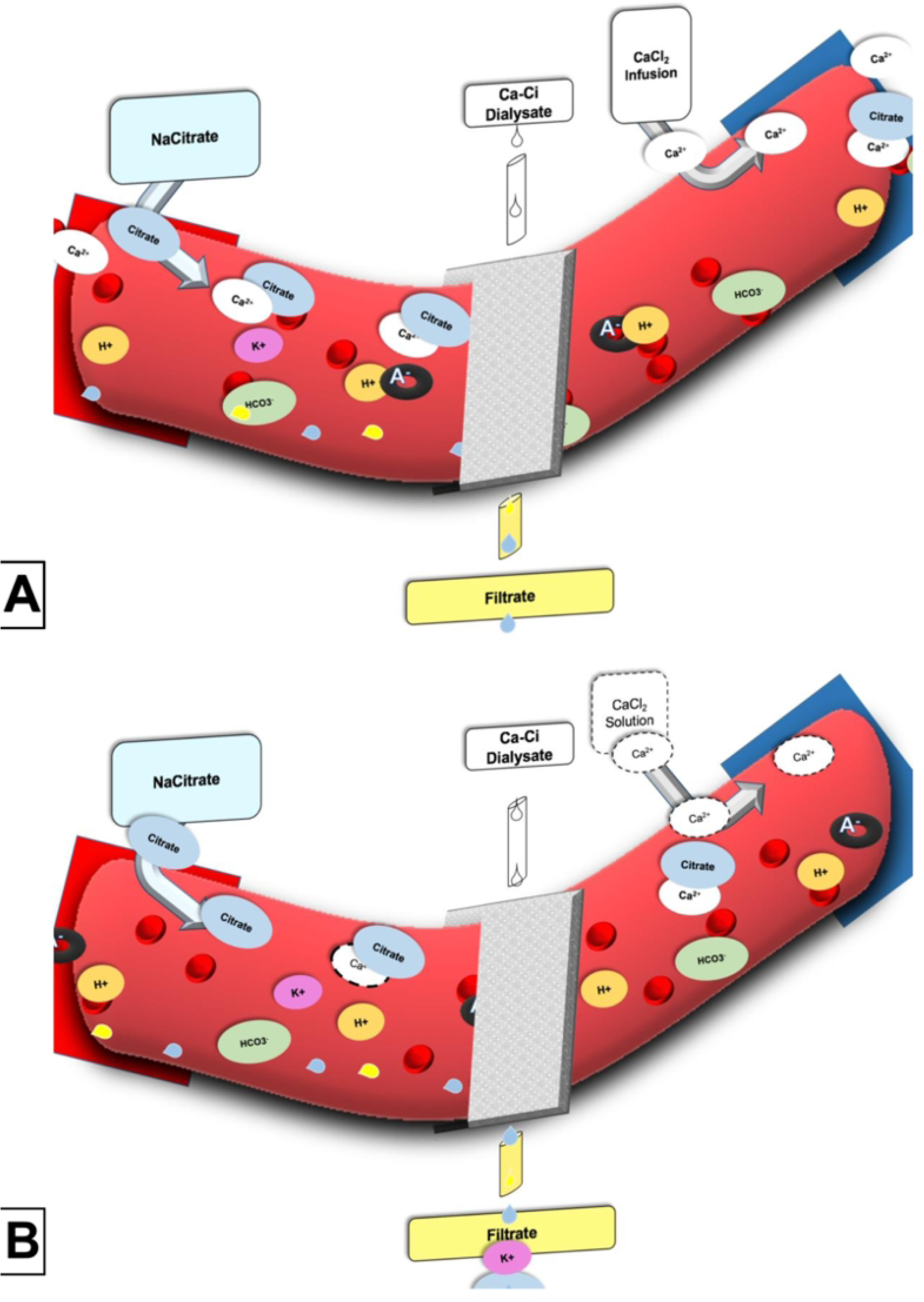
Visualization of the blood flow before and after the filter. *(A)* All parameters are within the target range. Blood flow is visualized by red blood cells moving through the system. Dialysate flow is depicted as transparent droplets falling from above the filter. The filter itself appears as a grey structure centrally positioned within the bloodstream. Citrate is infused from the left, binding to calcium ions flowing in proportion to the plasma calcium concentration. On the right, a calcium infusion is shown. The A-ring symbolizes the anion gap. Acid–base status is represented by hydrogen (H⁺) and bicarbonate (HCO₃⁻) ions. *(B)* Low plasma calcium concentration. This condition is indicated by dashed outlines around calcium ions and the absence of calcium near citrate molecules. The decision-support tool highlights an insufficient calcium infusion rate by displaying dashed lines around both the calcium infusion source and the infused calcium ions, prompting the user to increase the infusion rate.

This is the first study to evaluate the Visual Hemofilter, a novel visualization technology designed to support critical care professionals. The aim of this computer-based simulation study was to assess how the Visual Hemofilter influences decision-making during the adjustment of regional citrate anticoagulation, compared to conventional blood gas analysis with standard reference tables.

## Materials and Methods

This prospective, randomized, computer-based simulation study compared two approaches to managing regional citrate anticoagulation: the conventional method using blood gas analysis and reference tables versus decision support provided by the Visual Hemofilter. It was conducted across four intensive care units at the University Hospital Zurich. The study protocol was reviewed by the Ethics Committee of the Canton of Zurich, which issued a declaration of non-jurisdiction (BASEC-Nr. Req-2023-00415). Written informed consent was obtained from all participants, who were intensive care physicians and nurses. Participation was voluntary, with no financial compensation provided. Participants were randomly selected regardless of gender, age, professional role, or prior experience with regional citrate anticoagulation.

Participants were seated in a quiet room in front of a computer. After receiving a briefing on the study procedures, they provided written informed consent and completed a brief questionnaire to collect baseline characteristics. Responses were recorded using the online data collection tool *Harvest Your Data* (Wellington, New Zealand), accessed via various devices, including iPads (Apple Inc., Cupertino, CA, USA), Windows-based computers (Windows 11), and MacBook Air laptops (13”, macOS 10.15.7). All participants viewed a ten-minute instructional video on the Visual Hemofilter in German (S1 Appendix: English version; S2 Appendix: German version for reference), followed by a five-minute period for questions and review of specific video sections.

The trial consisted of two parts (Fig 2). In Part A, participants viewed 15 randomized video clips, each depicting a single parameter anomaly within the Visual Hemofilter (VHF) interface. Following each 45-second video, participants were asked to identify the anomaly from a list of 15 possible options (Table 1). Response time was unrestricted. This phase also served as a familiarization exercise, allowing participants to become accustomed to the new visualization technology

**Fig 2.**
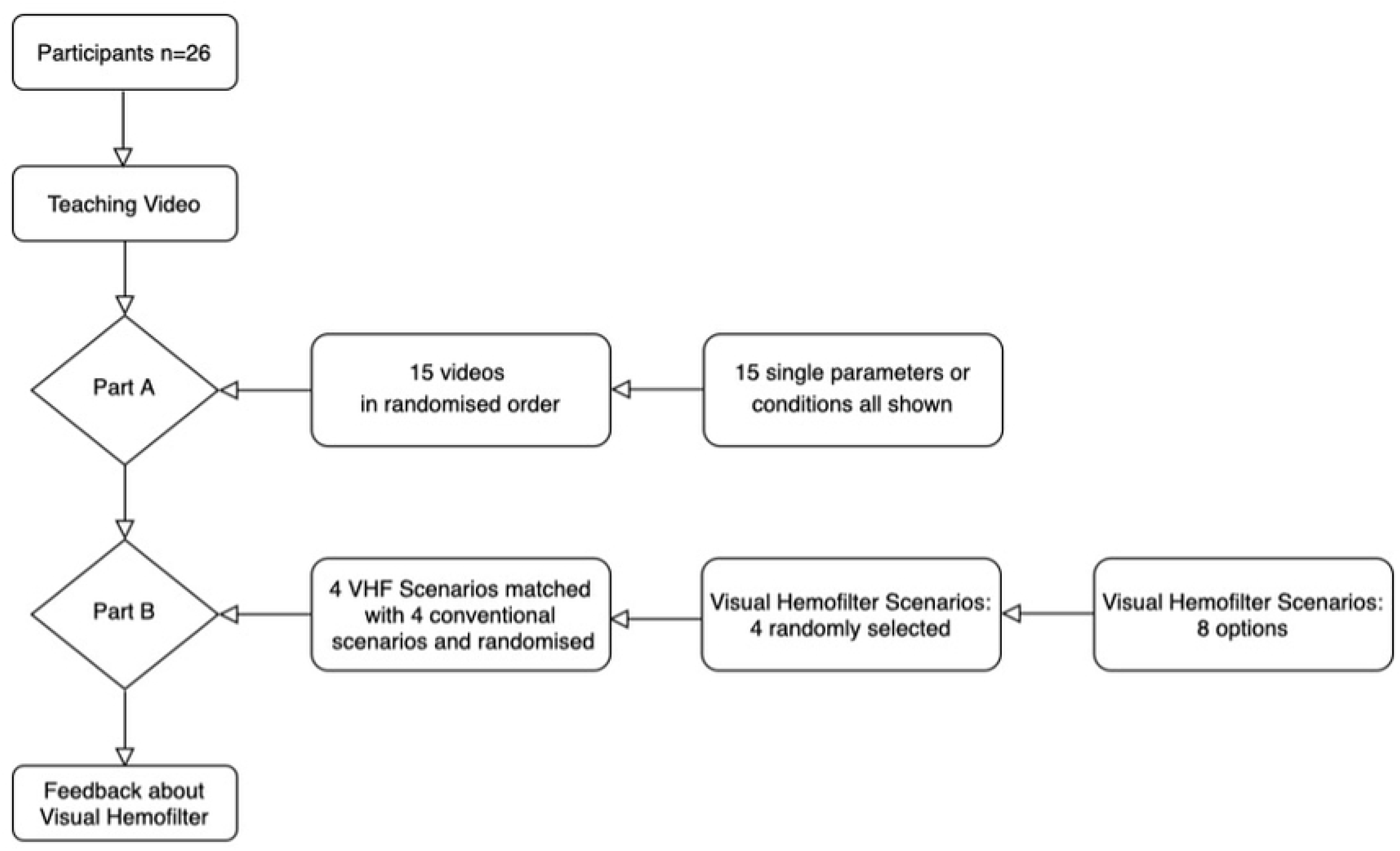
Study flow diagram. In Part A, participants viewed 15 video clips, each illustrating an anomaly in a single Visual Hemofilter (VHF) parameter. No conventional monitoring modalities were used during this phase. In Part B, participants completed four randomly selected scenarios using the Visual Hemofilter, each matched with an equivalent scenario using conventional blood gas analysis and reference tables. All scenarios were presented in randomized order.

**Table 1.** Part A Visual Hemofilter scenarios: single-parameter abnormalities. *Note:* In the case of citrate accumulation, high anion gap acidosis is accompanied by an increased need for calcium supplementation, as calcium remains bound to citrate due to impaired citrate metabolism. When calcium is adequately supplemented, systemic calcium levels may remain within range; however, the elevated substitution rate will still require clinical attention.

| Nr | Single status or condition | Anomaly |
| --- | --- | --- |
| 1 | Blood flow | too fast |
| 2 | Blood flow | too slow |
| 3 | Dialysate flow | too fast |
| 4 | Dialysate flow | too slow |
| 5 | Citrate infusion | too fast |
| 6 | Citrate infusion | too slow |
| 7 | Calcium infusion | too fast |
| 8 | Calcium infusion | too slow |
| 9 | Plasma calcium concentration | high |
| 10 | Plasma calcium concentration | low |
| 11 | Postfilter calcium concentration | high |
| 12 | Postfilter calcium concentration | low |
| 13 | Metabolic acidosis | Hydrogen ions high, Bicarbonate ions low |
| 14 | Metabolic alkalosis | Bicarbonate ions low, Hydrogen ions low |
| 15 | Citrate accumulation | Increased anion gap & low plasma calcium concentration |

Part B of the study involved the assessment of hemofilter setting adjustments (Fig 2). From a pool of eight possible scenarios, four were randomly selected for each participant (S3 Appendix). Each scenario presented a parameter anomaly along with the corresponding therapeutic adjustment (S4 Appendix). Every scenario (Table 2) was shown twice—once using the Visual Hemofilter (S4 Appendix) and once using conventional blood gas analysis results (S5 Appendix), accompanied by standard reference tables based on the CiCa-CVVHD system (Fresenius). All scenarios were presented in random order, each for 45 seconds. After each case, participants were asked: “What is the appropriate course of action in this case?” and selected from eight possible response options (Table 2).

**Table 2.**
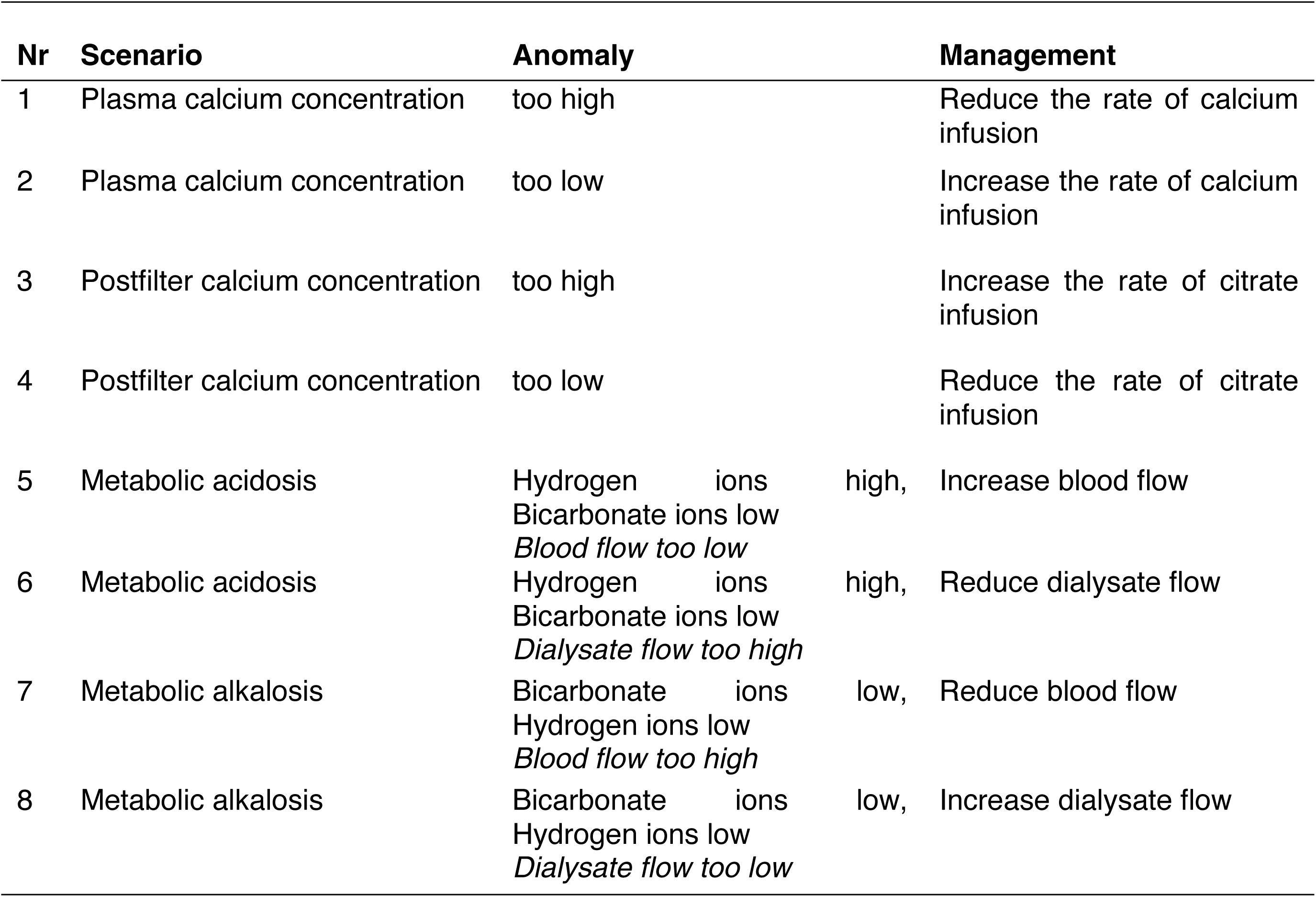
Complex Visual Hemofilter scenarios presented in Part B of the study.

The video or blood gas analysis display was terminated either 45 seconds after scenario onset or immediately upon participant response. Although each scenario was time-limited in presentation, participants had unlimited time to submit their answers. Response times were recorded using the online data collection tool *Harvest Your Data* (Wellington, New Zealand). Following each primary response, participants rated their diagnostic confidence and perceived cognitive workload. The sequence of scenarios in both study parts was randomized prior to the trial using *Research Randomizer*, Version 4.0. [19].

The primary outcome was decision accuracy, defined as the selection of the correct therapeutic adjustment for each scenario in Part B. In Visual Hemofilter scenarios, only one response was considered correct. In contrast, some conventional scenarios allowed for two correct options (e.g., in the case of metabolic acidosis, both increasing blood flow and reducing dialysate flow were acceptable). In such cases, selection of any correct response was counted as correct, provided no incorrect options were selected. All other therapy adjustments were classified as incorrect, except in predefined conventional scenarios where certain additional responses were considered acceptable but not required (e.g., in metabolic acidosis, selecting both “increase blood flow” and “increase citrate infusion rate” was not penalized, even if only one adjustment was necessary).

Time to decision was automatically recorded in seconds. After each scenario, participants rated their diagnostic confidence using a four-point Likert scale ranging from 1 (very unconfident) to 4 (very confident). Cognitive workload was assessed using the National Aeronautics and Space Administration Task Load Index (NASA-TLX), a validated instrument comprising six dimensions scored from 0 to 100. The “physical demand” dimension was excluded, as the study did not involve physically strenuous tasks, resulting in a modified five-item NASA-TLX with total scores ranging from 0 (lowest cognitive workload) to 100 (highest)[20]. In Part A of the study, the proportion of correctly identified anomalies was calculated for each scenario. To assess a potential learning effect, accuracy was analyzed in relation to the presentation order of scenarios for each participant, with the proportion of correct responses plotted against scenario sequence.

Descriptive statistics included means and standard deviations, as well as medians with interquartile ranges (IQR) for continuous variables, and frequencies with percentages for categorical variables. Poisson regression models were used to examine associations between the number of correctly recognized parameters per participant and factors such as professional role, age, gender, years of work experience, and years of experience with the hemofilter.

To assess the learning effect, we applied a mixed logistic regression model with a random intercept for each participant, using the order of video presentation as the independent variable. For the primary outcome analysis, we first conducted McNemar’s test, followed by a mixed logistic regression model with a random intercept per participant, adjusting for scenario.

Time to decision and cognitive workload were analyzed using linear mixed-effects models. Diagnostic confidence was analyzed by dichotomizing responses into “confident” versus “not confident,” and applying a mixed logistic regression model. All mixed models included a random intercept for participant and accounted for covariates such as modality and scenario.

Statistical analyses were performed using R (Version 4.0.5; R Foundation for Statistical Computing, Vienna, Austria), and figures were created using GraphPad PRISM (Version 8.1.1; GraphPad Software Inc., CA, USA). A p-value < 0.05 was considered statistically significant.

## Results

Between May 12 and June 12, 2023, a total of 26 intensive care physicians and nurses were recruited. Table 3 summarizes their baseline characteristics. All participants completed both parts of the study in full compliance with the study protocol (Fig 2). Results for the primary and secondary outcomes are presented in Fig 3.

**Fig 3.**
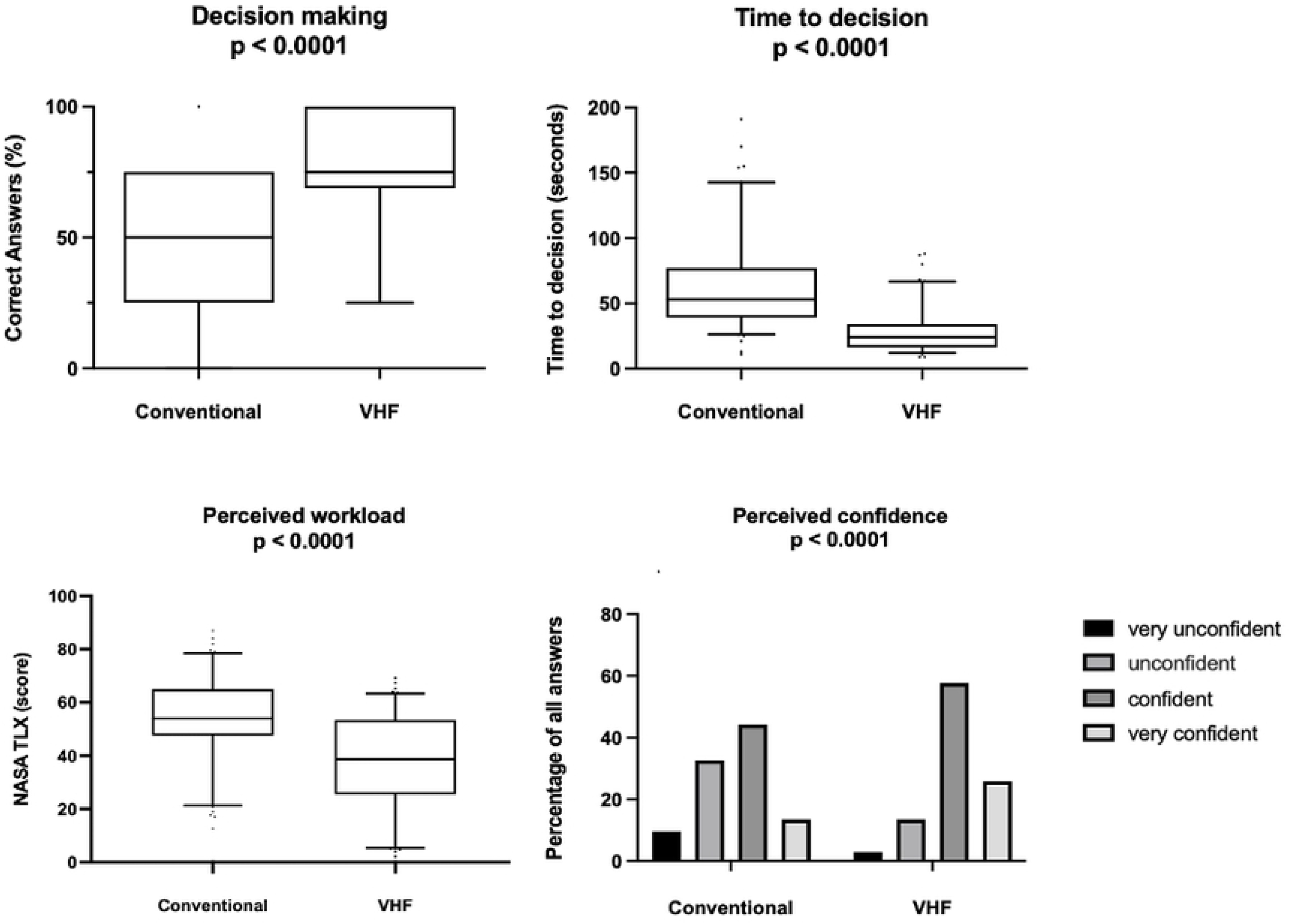
Group-level comparison between the conventional method and the Visual Hemofilter (VHF) (n = 26). Box plots display medians with interquartile ranges (IQR); whiskers represent the 5th to 95th percentiles, and individual dots indicate outliers. Perceived workload: Measured using the NASA Task Load Index (NASA-TLX; scale 0–100). Perceived confidence: Rated on a four-point scale (very unconfident, unconfident, confident, very confident); total number of responses per modality = 104. Statistical analysis was performed using mixed logistic regression models for correct responses and diagnostic confidence (dichotomized as confident vs. unconfident). Mixed linear models were used to analyze time to decision and perceived cognitive workload (NASA-TLX score).

**Table 3.** Participant and study characteristics. Values are presented as number (%), median (range), or mean (SD). *In Part B of the study, when the conventional modality was used for metabolic acidosis or alkalosis scenarios, any correct adjustment to blood flow or dialysate flow was considered a correct answer.

| Participant characteristics | Overall (N=26) |
| --- | --- |
| Gender; female | 17 (65.4%) |
| Age (years) |  |
| Mean (SD) | 39.0 (7.4) |
| Median [Min, Max] | 37 [27, 57] |
| Professional role |  |
| Senior physician (%) | 5 (19.2%) |
| Resident physician | 4 (15.4%) |
| Intensive care nurse | 16 (61.5%) |
| Intensive care nurse in training | 1 (3.8%) |
| Job experience (years) |  |
| Mean (SD) | 13.8 (7.8) |
| Median [Min, Max] | 14 [1, 31] |
| Experience with regional citrate anticoagulation (years) |  |
| Mean (SD) | 8.0 (5.5) |
| Median [Min, Max] | 7 [0, 23] |
| Study characteristics |  |
| Part A: 15 single anomaly videos shown as Visual Hemofilter | 390 |
| Part B: Computer-based simulations |  |
| Total | 208 |
| - displayed as Visual Hemofilter | 104 |
| - displayed as conventional scenarios using reference tables* | 104 |
| Number of times each scenario was shown using each modality |  |
| High plasma calcium concentration | 11 |
| Low plasma calcium concentration | 11 |
| High postfilter calcium concentration | 16 |
| Low postfilter calcium concentration | 12 |
| Metabolic acidosis - Increase blood flow | 16 |
| Metabolic acidosis - Reduce dialysate flow | 17 |
| Metabolic alkalosis - Reduce blood flow | 12 |
| Metabolic alkalosis - Increase dialysate flow | 9 |

### Decision-making

Use of the Visual Hemofilter significantly improved decision-making among intensive care professionals. The overall proportion of correct therapeutic decisions was 75% (95% CI: 64%–86%) with the Visual Hemofilter and 49% (95% CI: 38%–60%) with the conventional modality. McNemar’s test indicated a significant difference between the two methods, with an odds ratio of 3.45 (p = 0.0002) in favor of the Visual Hemofilter. These findings were further supported by a mixed logistic regression model, which yielded an odds ratio of 3.96 (95% CI: 2.03–7.73; p < 0.0001), indicating that participants were nearly four times more likely to make the correct therapeutic decision when using the Visual Hemofilter. An individual participant-level analysis revealed that 18 of 26 professionals performed better with the Visual Hemofilter, while an additional five participants performed equally well with both modalities (Fig 4).

**Fig 4.**
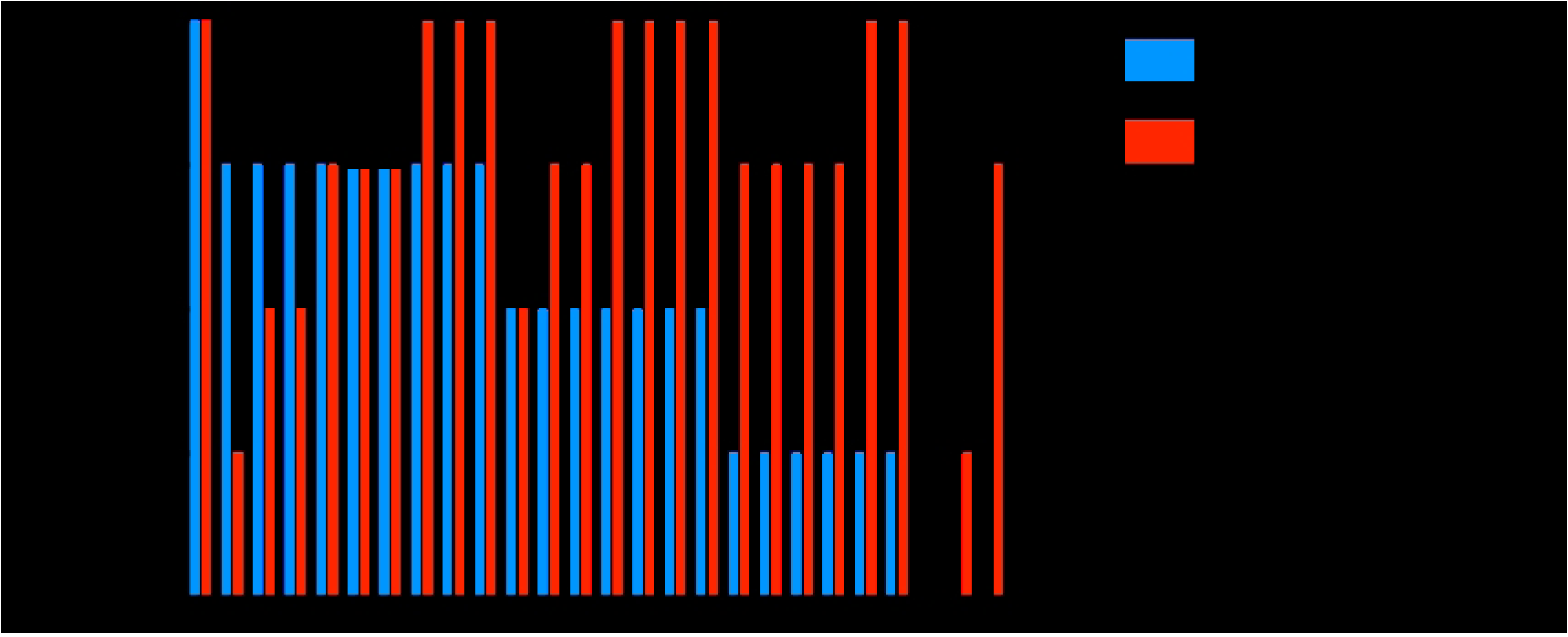
Decision-making at the individual participant level. Correct response rates for each of the 26 participants, ranked by the proportion of correct therapeutic decisions made using the Visual Hemofilter (VHF) and the conventional method.

### Time to Decision-Making

The mean time to reach a decision using the conventional method was 53 seconds (median: 61; IQR: 39–74; range: 11–191). In contrast, decision time decreased to a mean of 24 seconds with the Visual Hemofilter (VHF) (median: 28; IQR: 17–34; range: 9–88). This corresponds to a mean reduction of 33 seconds when using the VHF. A mixed linear regression model confirmed this time-saving benefit, yielding a coefficient of –33.3 seconds (95% CI: –39.4 to –27.2; p < 0.0001).

### Confidence in Decision-Making

In 27 out of 104 cases (26.0%), participants reported being “very confident” when using the Visual Hemofilter (VHF), compared to 14 cases (13.5%) with the conventional method. Additionally, 60 cases (57.7%) using the VHF were rated as “confident,” versus 46 cases (44.2%) with conventional tools. In contrast, participants reported being “unconfident” in 14 cases (13.5%) with the VHF and in 34 cases (32.7%) using the conventional method. “Very unconfident” ratings were given in only 3 cases (2.9%) with the VHF, compared to 10 cases (9.6%) with the conventional method. Overall, the odds of being confident in decision-making were 5.41 times higher when using the Visual Hemofilter (OR 5.41; 95% CI: 2.49–11.77; p < 0.0001).

### Cognitive Workload

The mean NASA Task Load Index (NASA-TLX) score for the conventional modality was 54.0 (median: 53.4; IQR: 47.8–64.6; range: 12.6–87.0). In contrast, the mean score significantly decreased to 38.6 with the Visual Hemofilter (VHF) (median: 38.2; IQR: 26.2–53.2; range: 2.2–69.2). A linear mixed-effects model confirmed a statistically significant reduction in perceived cognitive workload associated with VHF use, yielding a coefficient of –15.1 (95% CI: –18.9 to –11.1; p < 0.0001).

### Parameter Recognition

In Part A of the study, participants were presented with 15 single-parameter Visual Hemofilter (VHF) anomaly scenarios. The mean number of correctly recognized parameters was 11.3 (SD: 3.08), corresponding to a mean recognition rate of 75.6% (SD: 20.6%). The median number of correct identifications per participant was 12 (range: 3–15), equivalent to 80% accuracy (range: 20–100%). The percentage of correct parameter recognition for each scenario is summarized in Table 4.

**Table 4.** Proportion (%) of correctly recognized parameters by scenario (n = 26).

| Part A scenario | Proportion (%) of correctly recognized parameters / conditions in Part A |
| --- | --- |
| Blood flow too fast | 26/26 (100) |
| Calcium infusion too fast | 24/26 (92) |
| Citrate infusion too fast | 23/26 (89) |
| Metabolic alkalosis | 23/26 (89) |
| Blood flow too slow | 22/26 (85) |
| Dialysate flow too fast | 22/26 (85) |
| Metabolic acidosis | 22/26 (85) |
| Calcium infusion too slow | 21/26 (81) |
| Dialysate flow too slow | 21/26 (81) |
| Citrate accumulation | 18/26 (69) |
| Plasma [Calcium] too low | 17/26 (65) |
| Postfilter [Calcium] too high | 16/26 (62) |
| Citrate infusion too slow | 15/26 (58) |
| Plasma [Calcium] too high | 15/26 (58) |
| Postfilter [Calcium] too high | 10/26 (39) |

The study explored potential associations between parameter recognition and several covariates, including age, gender, professional role, overall job experience, and experience with regional citrate anticoagulation. However, univariate Poisson regression models revealed no statistically significant associations with any of these variables (p > 0.05).

### Learning Effect

A mixed logistic regression model indicated moderate evidence of a learning effect: with each successive scenario in Part A, the odds of correctly identifying the parameter increased by a factor of 1.06 (95% CI: 1.00–1.13; p = 0.047) (Fig 5). This finding suggests that participants’ ability to identify VHF parameters improved with repeated exposure, indicating a learning effect over time.

**Fig 5.**
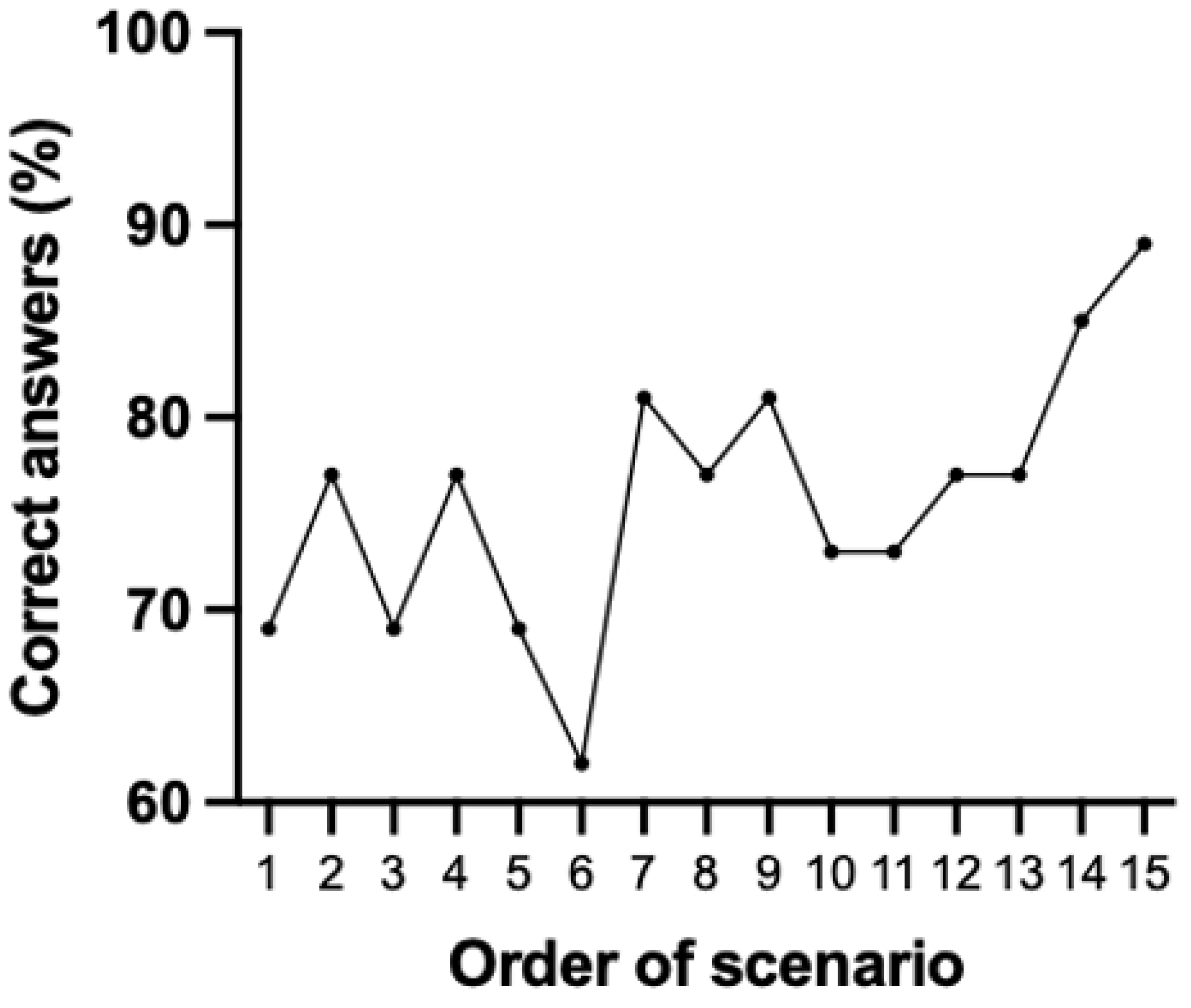
Proportion (%) of correct responses across all participants (N = 26), plotted by the presentation order of the 15 Visual Hemofilter scenarios in Part A.

## Discussion

The current landscape in critical care presents numerous challenges, including increasing patient volumes, rising case complexity [21], a global shortage of qualified staff, and heightened expectations for patient safety and quality of care [22–25]. These factors place a substantial burden on intensive care staff. As patient monitoring becomes more frequent and detailed, and as patient-to-caregiver ratios—particularly among nursing staff—increase, the interpretation of complex monitoring data contributes significantly to clinicians’ cognitive workload [26–28]. This increased burden can lead to cognitive fatigue and reduced vigilance, which may compromise patient safety, prolong hospital stays, and contribute to higher mortality rates [24, 29–34].

Acute kidney injury (AKI) affects 20–50% of intensive care unit (ICU) patients and is associated with increased mortality [21]. A considerable proportion of these patients require renal replacement therapy, often using regional citrate anticoagulation (RCA), which adds further to the clinical workload of ICU staff.

Our data suggest that the Visual Hemofilter has the potential to support critical care professionals in monitoring RCA and making related clinical decisions. These findings are consistent with prior research on other situation awareness–based, human-centered visualization technologies [1]. Notable examples include the Philips Visual Patient Avatar, a perioperative and critical care monitoring tool that translates vital signs into a single-indicator patient avatar[1–5, 35]; Visual Blood, which represents blood gas parameters and their physiological interactions as animated 3D icons flowing through a virtual vessel[1]; and Visual Clot, which visualizes viscoelastic test results (e.g., ROTEM®) as a dynamic 3D model of clot formation [36].

In addition to improving diagnostic accuracy and enabling faster decision-making, these visualization technologies have been shown to reduce perceived cognitive workload and increase diagnostic confidence among both medical and nursing staff [2–5]. These encouraging results highlight the potential of such tools to support healthcare professionals in enhancing patient care and outcomes within critical care environments. Visualization tools may be particularly valuable for junior staff, during night shifts, or in settings with limited personnel—circumstances in which opportunities for double-checking decisions are reduced. Moreover, clinicians who do not regularly manage RCA may benefit from the Visual Hemofilter both as a refresher on key concepts and as a real-time support tool for confirming therapeutic decisions.

These visualization technologies are grounded in a user-centered design approach influenced by the work of Endsley and Wittgenstein [17, 18], as well as key principles from human-computer interaction outlined in NASA’s publication by Degani et al. [37]. This design philosophy emphasizes the use of direct visual representations of data to enhance situation awareness. Wittgenstein’s notion of a “logical picture” suggests that an effective representation should mirror reality in a way that is immediately understandable , a logical picture should accurately reflect reality [18]. NASA’s guidance underscores the value of integrating all relevant information into a unified, well-organized display for optimal usability [37]. At their core, these technologies aim to improve interface design to support rapid, intuitive access to clinically relevant information, thereby strengthening caregivers’ situational awareness and decision-making [17].

Several limitations should be considered when interpreting our results. The study was conducted in a controlled test environment using simulated data, which may have amplified the observed effects. While we attempted to replicate some real-world ICU stressors—such as time constraints—by introducing a timed element, actual clinical environments often involve additional challenges, including frequent interruptions and multitasking. The study sessions took place in quiet rooms during daytime hours, with participants relieved of clinical duties, potentially underestimating the tool’s effectiveness under typical working conditions. Moreover, the scenarios were independent of real clinical cases, which may sometimes aid decision-making by providing contextual cues. Despite these limitations, the controlled study design allowed for a rigorous assessment of the Visual Hemofilter and represents the first formal evaluation of this novel tool in a research setting.

## Conclusion

This randomized, computer-based simulation study provides evidence that the Visual Hemofilter enhances the speed and accuracy of clinical decision-making, while also increasing user confidence and reducing cognitive workload. These findings suggest that the system may support critical care professionals by improving situational awareness and potentially reducing the risk of clinical error. Further validation in high-fidelity simulation environments and real-world clinical settings is warranted to confirm its effectiveness and generalizability.

## Data Availability

All relevant data underlying the findings of this study are publicly available. The supplementary video materials are available at Zenodo via the following DOI: https://doi.org/10.5281/zenodo.19552395). All other data are included within the manuscript and its Supporting Information files.

## List of abbreviations

AKI: Acute kidney injury
AGB: Arterial blood gas
CiCa-CVVHD: Citrate-based Continuous Veno-Venous Hemodialysis
CRRT: Continuous renal replacement therapy
Fig: Figure
ICU: Intensive care unit
IQR: Interquartile ranges
NASA-TLX: NASA Task Load Index
RCA: Regional citrate anticoagulation
SD: Standard deviation
VHF: Visual Hemofilter

## Acknowledgements

The authors are thankful to the study participants for their time and effort.

## Supporting information

## S1 Appendix: Visual Hemofilter teaching video in English

Supplementary videos are available at Zenodo (DOI: https://doi.org/10.5281/zenodo.19552395).

File name: Supplementary Video 1 - Visual Hemofilter Teaching Video in English File format: .mp4

Description: Blood flow is visualized by red blood cells moving through the system, while dialysate flow is depicted as transparent droplets falling above the filter. On the left side of the interface, a citrate infusion is shown; citrate binds to calcium ions, which are displayed in proportion to the plasma calcium concentration. A calcium infusion appears on the right side. The A-ring symbolizes the anion gap, and hydrogen (H⁺) and bicarbonate (HCO₃⁻) ions represent acid–base homeostasis. As the prototype was developed in German-speaking Switzerland, some interface labels are in German: *Citrat* (English:citrate); *Lösung* (solution); *Filtrat* (filtrate); *gebrauchtes Dialysat* (used dialysate).

## S2 Appendix: Visual Hemofilter teaching video in German

Supplementary videos are available at Zenodo (DOI: https://doi.org/10.5281/zenodo.19552395).

File name: Supplementary Video 2 - Visual Hemofilter Teaching Video German File format: .mp4

Description: Blood flow is symbolized by red blood cells, and dialysate flow by transparent droplets above the filter. On the left, a citrate infusion is shown; citrate binds to calcium ions, which flow proportionally to the plasma calcium ion concentration. A calcium infusion is displayed on the right. The A-ring symbolizes the anion gap, while hydrogen (H⁺) and bicarbonate (HCO₃⁻) ions indicate acid–base homeostatic status.

## S3 Appendix: Scenario coding

File name: S3 Appendix - Scenario coding File format: .docx

Description: Each scenario was coded to prevent pattern recognition. Fig 1: Visual Hemofilter scenarios depicting single-parameter anomalies used in Part A of the study. Scenario identifiers were anonymized using city names.

## S4 Appendix: VHF Scenario Snapshots

File name: S4 Appendix - VHF Scenario Snapshots File format: .docx

Description: Figures 1 to 9 show different Visual Hemofilter scenarios.

## S5 Appendix: VHF Blood gas analysis

File name: S5 Appendix -VHF Blood gas analysis File format: .pdf

Description: Examples of arterial and post-filter blood gas analyses used in the conventional modality during Part B of the study. Scenarios were anonymized using fruit names to prevent pattern recognition.

## Declarations

### Ethics Approval and Consent to Participate

The study protocol was reviewed by the Ethics Committee of the Canton of Zurich, Switzerland, which issued a declaration of non-jurisdiction (BASEC-Nr. Req-2023-00415). Written informed consent was obtained from all study participants.

### Consent for Publication

All participants provided written informed consent for publication of the collected data, with full anonymity maintained.

### Availability of Data and Materials

The datasets generated and analyzed during the current study are available from the corresponding author upon request.

### Competing Interests

J.B.L. is the first named inventor of Visual Hemofilter technology, for which the University of Zurich holds patent applications and design protections; potential royalties may follow successful commercialization. C.B.N. is an inventor of Visual Patient and Visual Patient Predictive technologies, for which the University of Zurich and Koninklijke Philips N.V. hold patents, patent applications, design protections, and trademarks. Joint development and licensing agreements exist with Philips Medizin Systeme Böblingen GmbH, Böblingen, Germany; Koninklijke Philips N.V., Amsterdam, The Netherlands; Philips Research/Philips Electronics Nederland BV, Eindhoven, The Netherlands; and Philips USA, Cambridge, MA, USA. Within the framework of these agreements, C.B.N. receives travel support, lecturing, and consulting honoraria and royalties. C.B.N. is an inventor of Visual Clot technology, with patent applications, design protections, and trademarks held by the University of Zurich. In case of successful commercialization, C.B.N. may receive royalties. C.B.N. is an inventor of Visual Blood technology, for which the University of Zurich holds patent applications and design protections; potential royalties may follow successful commercialization. C.B.N. received travel support, lecturing, and consulting honoraria from Instrumentation Laboratory—Werfen, Bedford, MA, USA. D.W.T. is the first named inventor of Visual Patient, Visual Patient Predictive, Visual Heart technologies, for which the University of Zurich and Koninklijke Philips N.V. hold patents, patent applications, design protections, and trademarks. Joint development and licensing agreements exist with Philips Medizin Systeme Böblingen GmbH, Böblingen, Germany; Koninklijke Philips N.V., Amsterdam, The Netherlands; Philips Research/Philips Electronics Nederland BV, Eindhoven, The Netherlands; and Philips USA, Cambridge, MA, USA. Within the framework of these agreements, D.W.T. receives research funding, travel support, lecturing, consulting honoraria and royalties.D.W.T. holds a position on the Philips Patient Safety Advisory Board. D.W.T. is the first named inventor of Visual Clot technology, with patent applications, design protections, and trademarks held by the University of Zurich. In the event of successful commercialization, D.W.T. may receive royalties. D.W.T. is the first named inventor of Visual Blood technology, for which the University of Zurich holds patent applications and design protections; potential royalties may follow successful commercialization. Additionally, D.W.T. received travel support, lecturing, and consulting honoraria from Instrumentation Laboratory—Werfen, Bedford, MA, USA, the Swiss Foundation for Anaesthesia Research in Zurich, Switzerland, and the International Symposium on Intensive Care and Emergency Medicine in Brussels, Belgium. D.W.T. is an inventor of Visual Hemofilter technology, with patent applications and design protections held by the University of Zurich. In case of successful commercialization, D.W.T. may receive royalties.

The other authors report no conflicts of interest regarding this paper.

## Funding

The Institute of Anesthesiology of the University Hospital of Zurich, Zurich, Switzerland and the University of Zurich, Zurich, Switzerland funded this project.

## Authors’ Contributions

Conceptualization: J.B.L., E.K., D.W.T.; Methodology: J.B.L., G.G., C.B.N., E.K., D.W.T.; Data Curation: J.B.L., G.G., B.R., J.W., H.S., P.M., M.H.; Formal Analysis: J.B. and J.B.L.; Investigation: J.B. and J.B.L.; Software: J.B.L.; Writing – Writing - original draft: J.B.L., G.G., D.W.T.; Writing - review & editing: J.B.L., G.G., B.R., J.B., J.W., H.S., C.B.N., P.M., M.H., E.K., D.W.T.; Visualization: J.B.L.; Supervision: J.B.L.; Project administration: J.B.L. All authors read and approved the final manuscript.

All authors have approved the submitted version (and any substantially revised version involving their contributions). They agree to be personally accountable for their own work and to help ensure that any questions about the accuracy or integrity of the entire work are appropriately investigated and resolved.

## Notes

### Competing Interest Statement

I have read the journal's policy and the authors of this manuscript have the following competing interests: J.B.L. is the first named inventor of Visual Hemofilter technology, for which the University of Zurich holds patent applications and design protections potential royalties may follow successful commercialization. C.B.N. is an inventor of Visual Patient and Visual Patient Predictive technologies, for which the University of Zurich and Koninklijke Philips N.V. hold patents, patent applications, design protections, and trademarks. Joint development and licensing agreements exist with Philips Medizin Systeme Böblingen GmbH, Böblingen, Germany Koninklijke Philips N.V., Amsterdam, The Netherlands Philips Research/Philips Electronics Nederland BV, Eindhoven, The Netherlands and Philips USA, Cambridge, MA, USA. Within the framework of these agreements, C.B.N. receives travel support, lecturing, and consulting honoraria and royalties. C.B.N. is an inventor of Visual Clot technology, with patent applications, design protections, and trademarks held by the University of Zurich. In case of successful commercialization, C.B.N. may receive royalties. C.B.N. is an inventor of Visual Blood technology, for which the University of Zurich holds patent applications and design protections potential royalties may follow successful commercialization. C.B.N. received travel support, lecturing, and consulting honoraria from Instrumentation Laboratory-Werfen, Bedford, MA, USA. D.W.T. is the first named inventor of Visual Patient, Visual Patient Predictive, Visual Heart technologies, for which the University of Zurich and Koninklijke Philips N.V. hold patents, patent applications, design protections, and trademarks. Joint development and licensing agreements exist with Philips Medizin Systeme Böblingen GmbH, Böblingen, Germany Koninklijke Philips N.V., Amsterdam, The Netherlands Philips Research/Philips Electronics Nederland BV, Eindhoven, The Netherlands and Philips USA, Cambridge, MA, USA. Within the framework of these agreements, D.W.T. receives research funding, travel support, lecturing, consulting honoraria and royalties. D.W.T. holds a position on the Philips Patient Safety Advisory Board. D.W.T. is the first named inventor of Visual Clot technology, with patent applications, design protections, and trademarks held by the University of Zurich. In the event of successful commercialization, D.W.T. may receive royalties. D.W.T. is the first named inventor of Visual Blood technology, for which the University of Zurich holds patent applications and design protections potential royalties may follow successful commercialization. Additionally, D.W.T. received travel support, lecturing, and consulting honoraria from Instrumentation Laboratory?Werfen, Bedford, MA, USA, the Swiss Foundation for Anaesthesia Research in Zurich, Switzerland, and the International Symposium on Intensive Care and Emergency Medicine in Brussels, Belgium. D.W.T. is an inventor of Visual Hemofilter technology, with patent applications and design protections held by the University of Zurich. In case of successful commercialization, D.W.T. may receive royalties. The other authors report no conflicts of interest regarding this paper.

### Funding Statement

Yes

### Author Declarations

The study protocol was reviewed by the Ethics Committee of the Canton of Zurich, Switzerland, which issued a declaration of non-jurisdiction (BASEC-Nr. Req-2023-00415).

